# Bioelectronic platform enables lectin-based enrichment of columnar cell clusters for Barrett’s oesophagus detection

**DOI:** 10.1101/2025.06.17.25328546

**Authors:** Suraj Pavagada, Neus Masque-Soler, Zixuan Lu, Ying Fu, Janire Saez, Ahsen Ustaoglu, Andrea Bistrovic Popov, John Lizhe Zhuang, Ioanna Mela, Ljiljana Fruk, Róisín M. Owens, Rebecca C. Fitzgerald

**Affiliations:** Early Cancer Institute, Department of Oncology, University of Cambridge, Cambridge, UK; Department of Chemical Engineering & Biotechnology, University of Cambridge, Cambridge, UK; Pure and Applied Chemistry, University of Strathclyde, Glasgow, UK; Department of Zoology and Animal Cell Biology, Faculty of Pharmacy, University of the Basque Country (UPV/EHU), Basque Country, Spain; Basque Foundation for Science, IKERBASQUE, Bilbao, Spain; Department of Pharmacology, University of Cambridge

## Abstract

Enriching diagnostically relevant cells from heterogeneous clinical samples is critical for enabling accurate detection and molecular analysis. Bioelectronic platforms offer a promising approach to this challenge by combining selective capture with real-time, label-free monitoring. We focus on Barrett’s oesophagus (BE), a precursor to oesophageal adenocarcinoma (EAC), where current non-endoscopic tools like the capsule-sponge yield samples dominated by background squamous cells, limiting diagnostic sensitivity. We present a bioelectronic enrichment platform that selectively captures and thermally releases columnar cells from capsule-sponge samples. The system employs a ring microelectrode array functionalised with the lectin ECA—identified here as a selective marker of columnar cells in BE—and coated with a thermo-responsive PEDOT-pNIPAAam polymer. Capture and detachment of cells were monitored using both electrochemical impedance and optical imaging, enabling real-time, label-free feedback. Applied to clinical samples, our platform enriched viable columnar cells, enhancing downstream molecular readouts. This approach integrates seamlessly with non-invasive sampling workflows and expands the utility of bioelectronic tools for early cancer diagnostics.

## Introduction

The ability to enrich specific cell populations from heterogeneous clinical samples is critical for improving diagnostic accuracy, enabling downstream molecular analyses and advancing point-of-care technologies^1,2^. This need spans numerous biomedical contexts, including early cancer detection, where diagnostically relevant cells are often outnumbered by background populations^3,4^. Conventional enrichment methods such as FACS or MACS, while effective in controlled lab settings, are often impractical for clinical workflows due to equipment cost, cell aggregation issues, and incompatibility with downstream diagnostics^5^.

Bioelectronic microfluidic platforms offer an emerging alternative, enabling label-free, selective cell capture with integrated real-time monitoring^6–10^. Functionalised with molecular recognition elements such as antibodies or lectins, these platforms can dynamically control capture and release, while electrochemical impedance spectroscopy (EIS) enables non-destructive readout^11–14^. However, their clinical deployment remains limited, particularly for complex, unprocessed samples where high specificity and robustness are essential^15–17^.

We focus on Barrett’s oesophagus (BE)—a pre-malignant condition where the squamous oesophageal epithelium is replaced with columnar cells^18^. BE significantly increases the risk of oesophageal adenocarcinoma (EAC), a cancer with <20% five-year survival when detected at Stage 3 or beyond (Fig. 1a)^19^. Longer BE segments correlate with higher risk of progression to high-grade dysplasia or EAC, making early detection critical^20^. Minimally invasive tools such as the capsule-sponge have enabled broader screening, retrieving oesophageal cells without the need for endoscopy^21^. When coupled with biomarker tests like trefoil factor 3 (TFF3), this approach has improved BE detection^22,23^. However, capsule-sponge samples are dominated by squamous cells—often >90%— which poses challenges for additional biomarker or molecular analyses (Fig. 1b)^23–26^.

**Figure 1.**
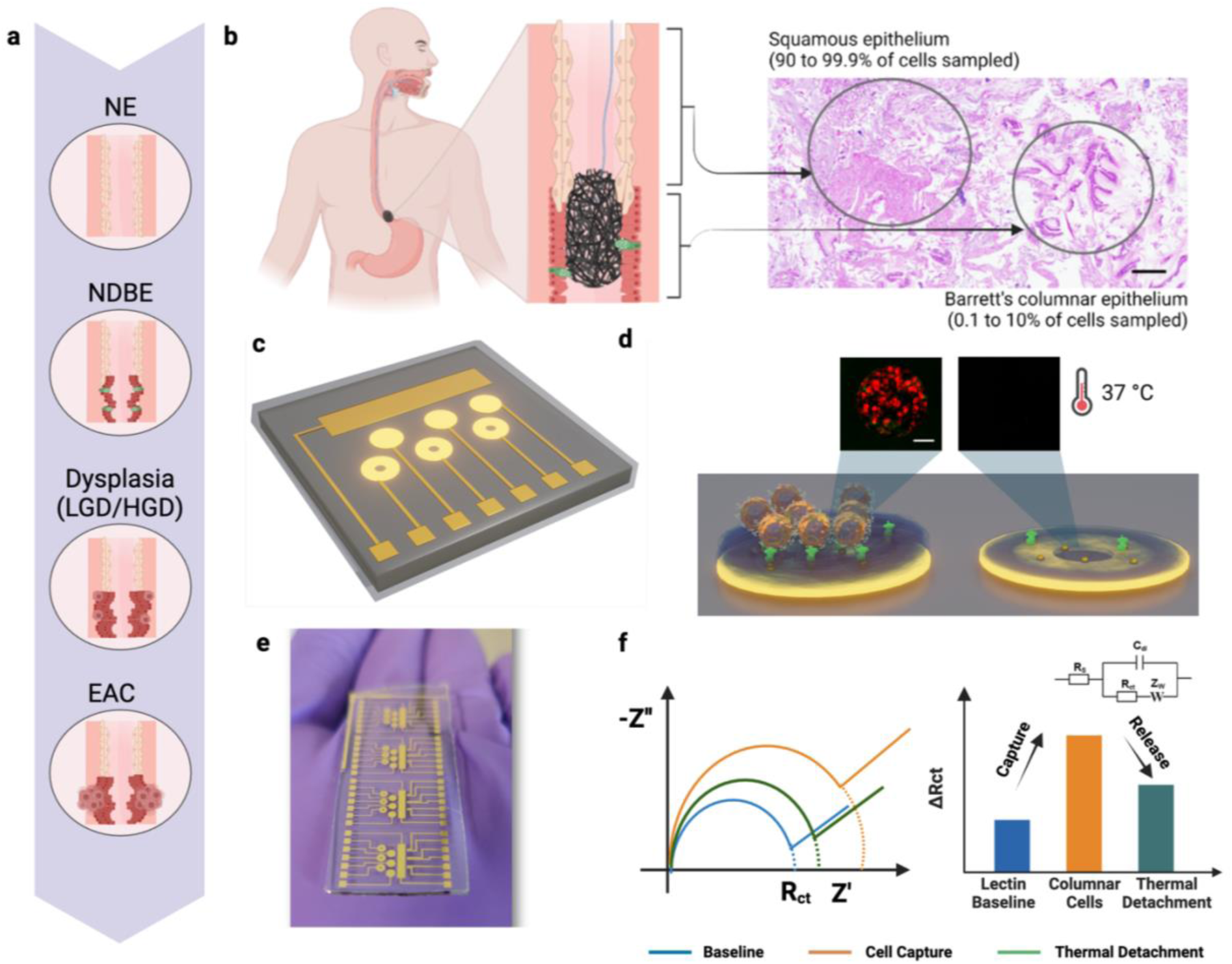
Bioelectronic enrichment platform for enrichment of columnar cells from capsule-sponge sampling. a) Transformation of normal squamous oesophagus (NE) to columnar lined non-dysplastic BE, and progression to stages of dysplasia (LGD/HGD) and EAC. b) Capsule-sponge sampling of the gastro-oesophageal junction. A H&E-stained slide pathology image of the processed cellular material from capsule-sponge depicting the two main types of cells collected. c) Bespoke fabricated gold electrode array pattern to facilitate simultaneous optical-electrochemical monitoring. d) Polymer coated electrodes facilitating lectin-based cell capture and release by thermal actuation. Inset depicting optical fluorescence visualisation of centre of the ‘ring’ electrode. e) Picture of a single fabricated chip consisting of four arrays. f) Electrochemical impedance spectroscopy (EIS)-based monitoring of cells captured on the polymer-coated electrode and after thermal release. Representative Nyquist plot (left) and bar plot (right) illustrating the quantification of electrochemical parameters based on the Randle’s equivalent circuit, including Rs (solution resistance), Rct (charge transfer resistance), Cdl (double-layer capacitance), and Z_W_ (Warburg impedance).

Lectins—proteins that bind to specific carbohydrate molecules on cell surfaces—have emerged as a promising class of cancer biomarkers^27,28^. Lectins offer a cost-effective alternative to mass spectrometry-based techniques and antibody use ^29,30^. Lectins have been shown to be of relevance in EAC, where specific cell-surface glycans are diminished as the disease progresses^31^. Expanding the use of lectin biomarkers in this context could establish them as valuable tools for evaluating cell-surface glycoproteins as markers for specific enrichment of BE and dysplastic cells.

Here, we present a bioelectronic lectin-functionalised platform that enables selective capture and thermal release of columnar cells from minimally invasive BE samples. We identified ECA as a lectin that selectively binds to columnar cells across the BE progression spectrum (Fig. 1a–c). The platform integrates a ring microelectrode array functionalised with ECA (Fig. 1d), coated with a thermo-responsive PEDOT-pNIPAAam layer (Fig. 1e–f), and designed to allow real-time electrochemical and optical monitoring of cell capture (Fig. 1g–h). When applied to clinical capsule-sponge samples, the system achieved efficient enrichment of viable columnar cells from heterogeneous populations. This enhanced the specificity and quality of downstream molecular analyses and holds promise as an integrable, label-free tool for non-endoscopic screening. Together, these results position the platform as a broadly applicable approach for selective cell enrichment in early cancer diagnostics.

## Results

### ECA lectin demonstrates selective binding to columnar cells across BE progression

Immunofluorescence (IF) staining was performed using a panel of 20 lectins, a subset of the lectin array examined in our previous study, selected based on their diverse glycan binding specificities, including O-glycan (LacNac, GalNac), sialic acid, fucose, and complex N-glycan binding classes, to comprehensively evaluate contextual tissue binding patterns^31,32^(ESI table 1,2). Lectins *Cytisus scoparius* lectin (CSA) and Peanut agglutinin (PNA) exhibited exclusive staining in gastric cardia while the lectin *Erythrina crista-galli* (ECA) exhibited strong binding to columnar epithelium of both gastric cardia (GC) and BE (ESI Fig. 1). In addition to ECA, four other lectins (GS-1, MAL-1) also demonstrated specific binding to both GC and BE tissues (ESI Fig.2). However, only lectin ECA exhibited binding to intestinal-type goblet cells, which are characteristic of BE tissue. As a result, ECA was selected for further evaluation in an independent cohort of clinical samples.

The IF staining pattern of lectin ECA was further tested in a total of 45 formalin fixed paraffin embedded (FFPE) biopsy tissues (n=15 each of normal squamous (NE), gastric cardia (GC) and BE) since these are the relevant cell types retrieved by non-endoscopic capsule-sponge devices on their removal from the stomach (Fig. 2a). Signal intensities were quantified as percentage area positivity, and indicated an exclusively columnar specific binding in all samples, i.e. all 15 NE tissues exhibited no ECA staining, while all other tissues showed strong epithelial columnar staining, including goblet cells in BE tissues. Representative images of IF staining patterns of FFPE biopsies and their quantifications are depicted in Fig. 2b.

**Figure 2.**
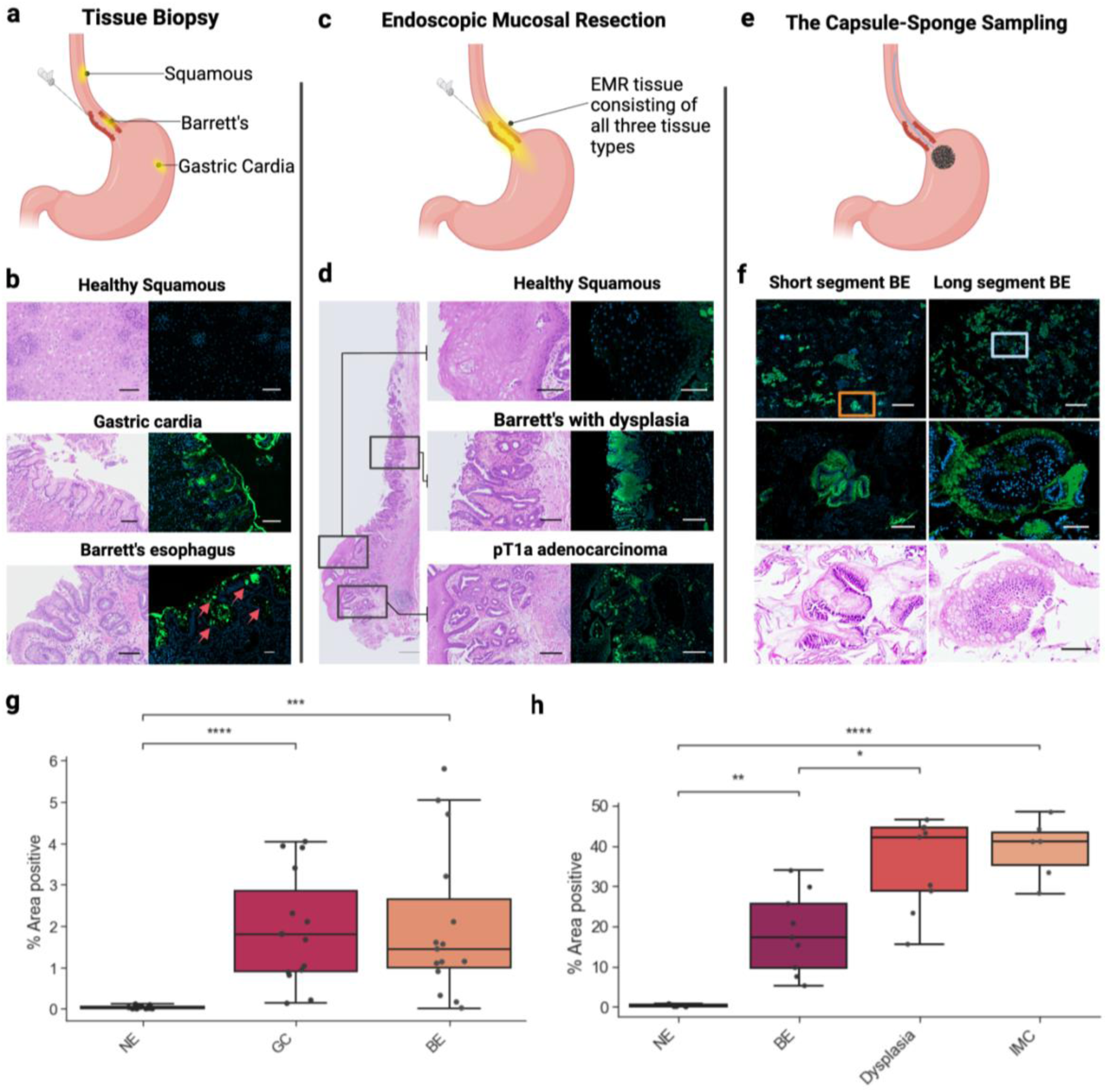
Lectin ECA validation as a cell surface marker for assessing the progression of normal oesophagus to BE through dysplasia and EAC in situ. a), c), e) Schematic of the biopsy trios, Endoscopic Mucosal Resection (EMR) and capsule-sponge samples collected for biomarker validation respectively. b), d), f) Representative adjacent H&E and IF magnified images with scale bars -200μm. Red arrows indicate goblet cells of IM. g), h) Quantified % area of positive staining in biopsy trios (n=45 regions of interest, ROI in n=15 samples) and EMR tissue (n=90 ROIs in n=12 samples). Pairwise independent t-test with Bonferroni correction, *** p ≤0.001, **** p ≤0.0001. NE-Normal Oesophagus (Squamous), GC – Gastric Cardia, BE – Barrett’s Oesophagus, IMC – Intramucosal Cancer.

Next, we investigated whether the binding of ECA is sustained in dysplastic BE and intramucosal cancer (IMC) tissues, or whether it decreases as reported in previous studies since it is important to be able to detect tissues transitioning to cancer. Endoscopic Mucosal Resections (EMR) can represent multiple stages of the disease, as they can contain several lesions at different stages, making them ideal for assessing ECA binding changes in early neoplasia. Immunofluorescence staining of EMR samples with ECA (Fig. 2c, d) showed a sustained increase in signal from BE as it progresses to dysplasia and IMC (Fig. 2h).

A competitive binding assay was conducted to determine the specific sugar residue that mediates binding between ECA lectin and BE/dysplastic tissue^33,34^. Lactose, a disaccharide composed of glucose and galactose linked by a β-1,4-glycosidic bond, and its two hydrolysis products (glucose and galactose) were used in competition assays with ECA lectin to determine binding specificity^35,36^. The results demonstrated that β-lactose significantly inhibited ECA binding, while its constituent monosaccharides glucose and galactose showed minimal competitive inhibition. This finding indicates that BE tissues predominantly express β-lactose moieties on their surface, a characteristic that persists in both dysplastic tissue and adenocarcinoma. (See ESI Fig. 3).

Lectin ECA staining was further tested on sections of capsule-sponge samples of patients (n=10) with known BE. We wanted to ensure that the lectin binding specificity is maintained in a heterogenous sampling consisting of several potential confounders including normal respiratory epithelium, and in some instances bacterial/fungal cells. Staining results indicated a remarkable specificity toward columnar cell groups, which was maintained throughout all samples tested (Fig. 2e, f). Quantification of the fluorescent signal, and assessment of stained cellular morphology, further confirmed the specificity of ECA for columnar cells amidst a vast background of squamous and other potentially confounding cells (ESI Fig. 4).

### PEDOT-pNIPAAM as a hybrid conductive-thermoresponsive polymer & ring microelectrode array fabrication

Having identified a potential marker for columnar cells, we sought to develop a system for enrichment of these cellular clusters. To enable point of care detection of BE and dysplasia, an electrochemical method amenable to portable operation was selected.

A microelectrode array consisting of a ring-electrode design was fabricated as per the parylene lift-off protocol previously stated^37^. The design includes a circular transparent window at its centre (ESI Fig. 6). Electrochemical assessment of these electrodes exhibited excellent robustness and homogeneity in baseline measurements, with a charge transfer resistance (Rct) of 920 Ω derived from EIS data, showing no significant differences to the standard circular electrode without the optically transparent window (ESI Fig. 7). To achieve selective capture and subsequent retrieval of the cells of interest, a thin-film coating of the smart polymer (PEDOT-pNIPAAM) along with lectin-functionalised AuNP was employed which showed greater homogeneity and sensitivity in specific cell capture over direct adsorption of lectins (ESI Fig. 8,9,10).

The polymer coated microelectrode array was functionalised with the lectin ECA through a series of steps to facilitate selective binding of columnar cell populations (Fig. 3a). The polymer characterisation upon AuNP integration and lectin attachment was carried out via Scanning Electron Microscopy (SEM) and Atomic Force Microscopy (AFM) (Fig. 3b). Fig. 3c,d shows the effect of each functionalisation stage on EIS measurements. An initial increase in ΔRct of 1.9kΩ was observed after spin-coating the hybrid polymer, likely due to the non-conductive pNIPAAM component. Following AuNP-lipoic acid incorporation, ΔRct decreased significantly by 2.4kΩ, reflecting enhanced conductivity. Subsequent lectin coupling via EDC-NHS chemistry led to an increase in ΔRct by 2.4kΩ, indicating the electrode’s sensitivity to biomolecule binding. Finally, cell capture caused a substantial rise in ΔRct of 16.4kΩ, attributed to surface blocking by the cell monolayer. The cell line TIF LifeAct-RFP was used to evaluate cell capture on lectin-functionalised polymer which was identified to be specific to the ECA lectin^38^. A one-way ANOVA confirmed statistically significant differences between all subsequently tested groups

**Figure 3.**
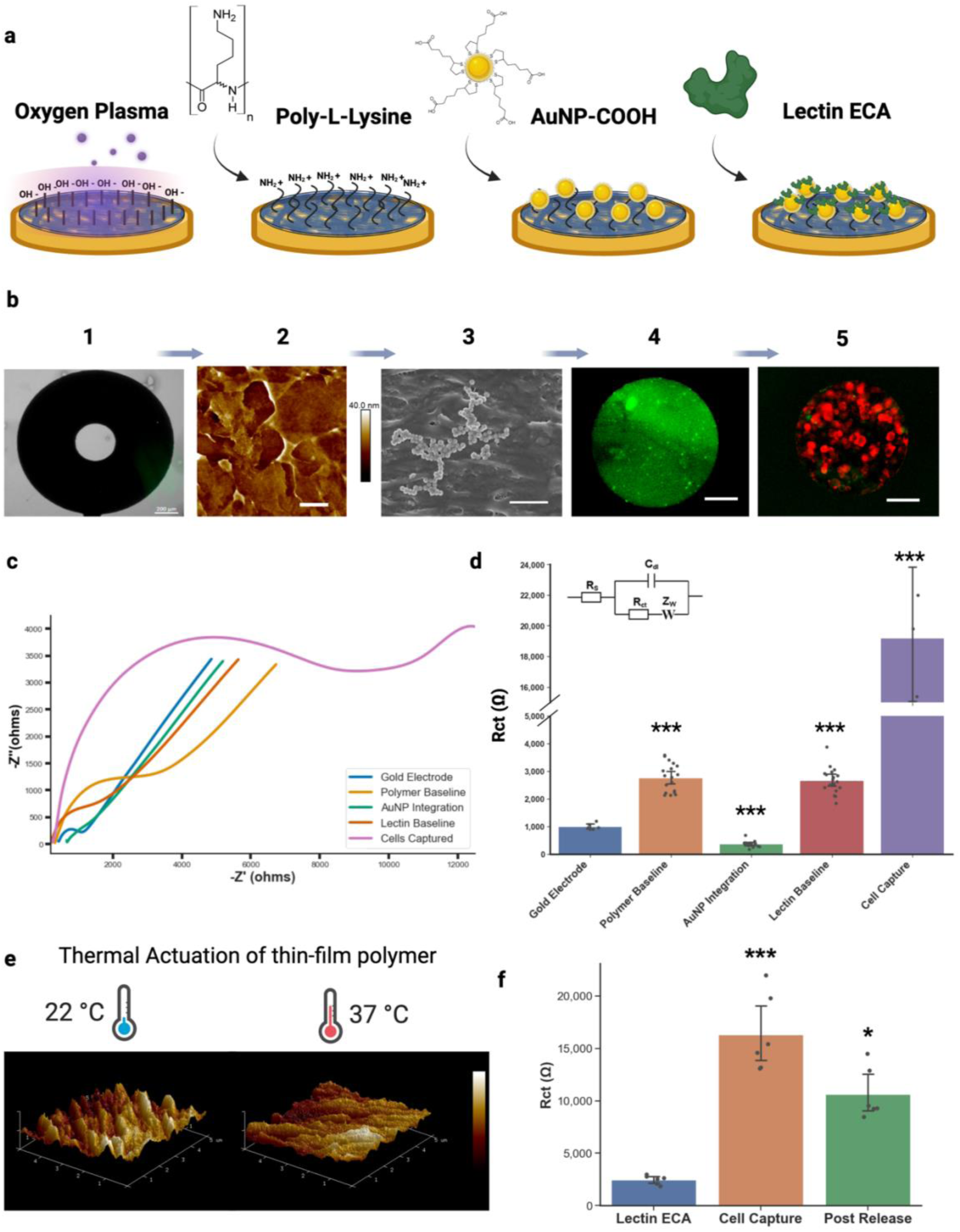
Functionalisation strategy for covalent coupling of lectin ECA onto the polymer-coated MEA. a) Schematic of each step in the functionalisation protocol. b) Graphical representation of each step followed by characterisation studies employed to track each layer of functionalisation. 1-Bare gold electrode and brightfield microscopy with scale bar -200μm, 2- polymer-coated electrode and AFM with scale bar - 1μm, 3- AuNP loading and SEM with scale bar – 500nm, 4- Lectin ECA-FITC coupling and fluorescence microscopy and 5- Fluorescence microscopy of TIF-lifeact-RFP cells captured on the lectin functionalised ring electrode, scale bar 200μm. c), d) Represent the EIS measurements taken during each step of the functionalisation process with the Nyquist plot of raw data and the simplified Randle’s circuit-based quantification, respectively. Error bars correspond to the standard deviation from the mean for n=18. Pairwise t-test with Bonferroni correction indicated statistically significant differences (*** p ≤0.001) for all conditions relative to its previous measurement. e) 3D rendering of AFM measurement of the hybrid polymer at room temperature, and after placing the polymer above the LCST at 37**°**C. Images correspond to a 5 µm × 5 µm scan area. The height color bar represents a vertical range of 0 to 30nm f) Simplified Randle’s circuit quantification of the EIS measurements for lectin baseline, cells captured at RT and released post thermal treatment of 37**°**C.

The effect of temperature on the thin-film hybrid polymer was assessed using AFM, which revealed a transition from a hydrophilic pillar network at room temperature to a flat hydrophobic surface above the polymer’s lowest critical stimulating temperature (LCST) at 37°C (Fig. 3e). This morphological shift influenced polymer-cell interactions, as cells bound to the polymer at room temperature were released at 37°C, with changes monitored both electrochemically via EIS and optically through fluorescence microscopy of the ring electrode (Fig.3f). Consistent with the data above, a significant increase in impedance was observed upon cells binding to the lectin-functionalised electrodes with a ΔRct of 17kΩ, followed by a drop in ΔRct to 10kΩ upon thermal actuation to release the cells. To further characterise the mechanism of cell release, we tested the effect of temperature after each of the functionalisation steps. EIS results confirmed that the polymer maintained its functionalisation with AuNP and lectins after LCST treatment (ESI Fig. 11).

### Static platform performance and dynamic columnar cell enrichment of capsule-sponge sample in flow

We first validated our platform in a static configuration, where functionalised electrodes were housed in a stationary well. The EIS response showed a linear dynamic sensing range between 200-40,000 cells per electrode with a detection limit of 140 cells (Fig. 4a). The thermal detachment efficiency across the array was 78% of the initial loading, with a retrieval rate of 95% cell viability as confirmed by the ViCell counter (ESI Fig. 12). To assess specificity, we conducted spike-in experiments using normal squamous oesophageal cells and TIF cells (ECA-specific) at 5% and 25% ratios. While pure squamous samples showed no binding, spiked samples exhibited ΔRct increases of 1.5kΩ and 14kΩ respectively, confirming selective capture of target cells within heterogeneous populations (Fig. 4b).This confirmed the lectin-functionalised electrodes’ ability to selectively capture target cells within a heterogeneous cell population.

**Figure 4.**
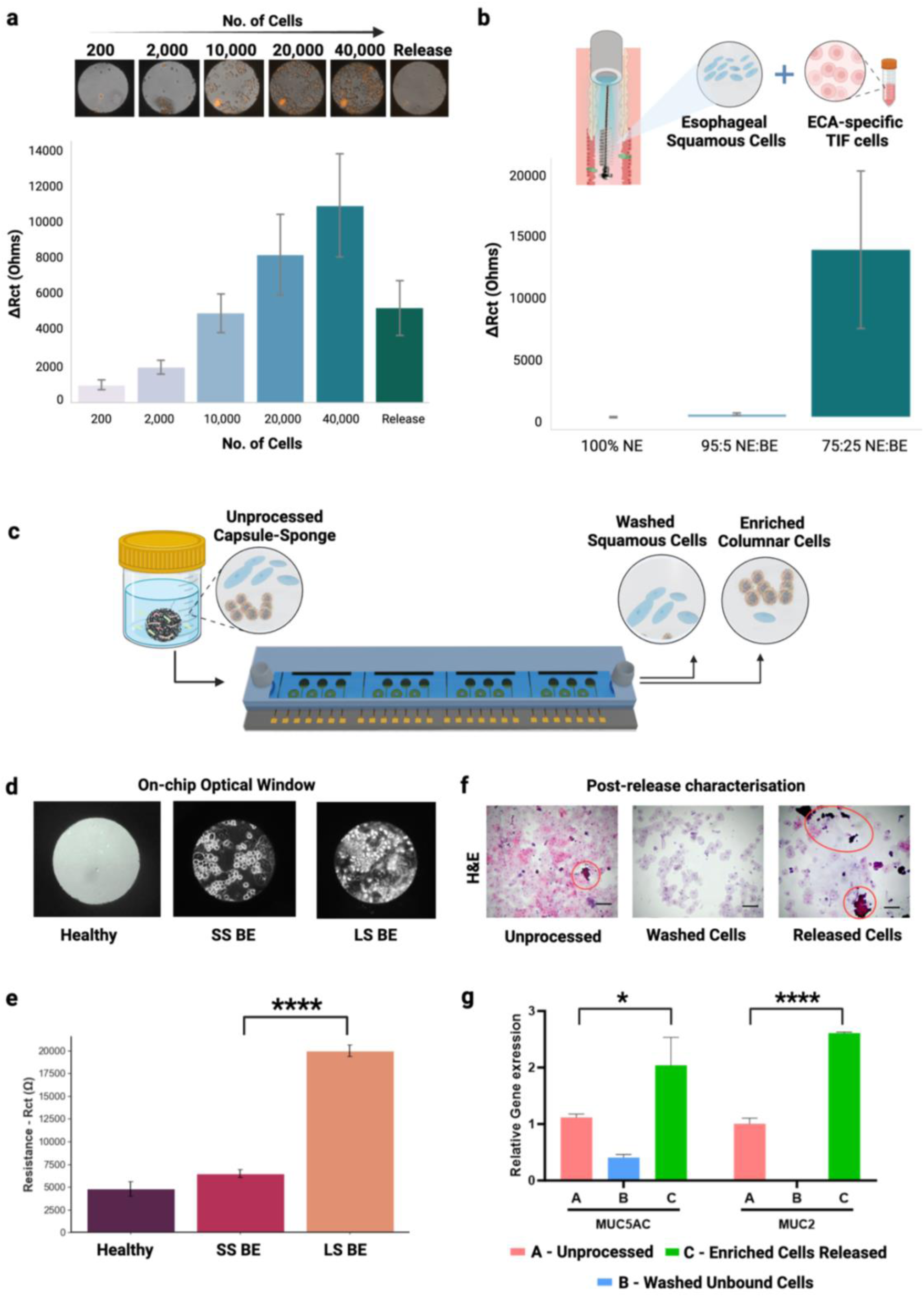
Bioelectronic Enrichment platform performance. a) EIS quantification demonstrating sensitivity and homogeneity of cell capture across three MEA chips (n=18). Representative images of optical monitoring of cell capture within the optical window is depicted above their respective concentration measurement. b) Rct quantification of EIS measurements indicating the selectivity of target cell capture in mixed sampling. Error bars correspond to the standard deviation from the mean for n=6 electrodes. Pairwise t-test with Bonferroni correction was carried out. Both ratios with TIFs spiked-in indicated a significant difference relative to the lectin baseline *p<0.05, *** p ≤0.001. c) Schematic of the microfluidic integrated bioelectronic platform used to facilitate dynamic enrichment of columnar cells from unprocessed capsule-sponge samples in flow. d) On-chip monitoring of columnar cell captured using optical brightfield microscopy after 1hr of sample processing e) Corresponding relative quantification of EIS measurements taken after 1hr of sample processing. A statistical t-test was carried out., and long segment BE case indicated a significant difference relative to the healthy volunteer/short segment BE case *** p ≤0.001. f)Downstream H&E staining of bound cells released from the platform compared to unprocessed sample g) RT-qPCR-based analysis of cells released from the platform by thermal actuation.

Building on these results, we extended the platform’s capability to a dynamic setting by testing it with multiple arrays to demonstrate dynamic cell enrichment from non-invasive capsule-sponge samples. Compared to static wells, microfluidic systems enhances cell capture efficiency by enabling controlled sample flow, and ensuring uniform exposure to the receptor-functionalized electrodes^39^. Oesophageal cytology sampling presents challenges due to its viscous mucosal layers and resulting large cell clumps, which can hinder effective contact between target cells and the functionalised surface. To overcome this, we utilized a microfluidic flow cell linking four arrays, facilitating more efficient cell capture across a larger area of functionalized surfaces. Patient capsule-sponge samples with clinically representative, varying lengths of BE and no BE were tested to validate the microfluidic enrichment platform.

After processing samples through the fluidic channel for 1 hour, on-chip brightfield microscopy of the optical window showed the extent of cells bound to the lectin-coated polymer surface of the electrodes (Fig. 4d). Corresponding EIS measurements revealed that impedance signals correlated with BE segment length (Fig. 4e). The ΔRct values were highest for long-segment BE (18.2kΩ), followed by short-segment BE (4.5kΩ) and healthy volunteer samples (1.5kΩ).

After thermal release, we collected three cellular fractions: (1) unprocessed capsule-sponge samples containing a heterogeneous mix of cells, (2) unbound cells, primarily squamous, and (3) target cells released from the platform, enriched for columnar cells. H&E staining showed that the enriched fraction consisted mainly of columnar cell clusters, with substantially reduced squamous cell contamination compared to the unprocessed samples (Fig. 4f). To verify the identity of lectin-bound cells captured on the electrode, we performed MUC5AC immunofluorescence staining on cells processed from the same sample (long segment BE) for 1 hour using a separate microfluidic chip. This confirmed the columnar phenotype of the captured cells (ESI Fig.13).

Assessment of gene expression by RT-qPCR was conducted to characterise the cell group enrichment from the capsule-sponge sampling. The extracted RNA samples from the sample aliquots were analysed by RT-qPCR using two mucin genes: *MUC5AC*, primarily expressed in gastric cardia columnar epithelium, and *MUC2*, for IM cells BE (Fig. 4g). There was a statistically significant difference in the expression of both *MUC5AC* (p value < 0.05) and *MUC2* (p value < 0.0001) between the enriched sample compared to the unprocessed capsule-sponge sampling.

## Discussion

This study presents a bioelectronic enrichment platform that enables selective capture, thermal release, and characterization of columnar cells from heterogeneous oesophageal samples, including those collected via the non-invasive capsule-sponge. By integrating lectin-based targeting with nanostructured, thermo-responsive conducting polymer coatings, we achieved efficient enrichment of viable cells. These included diagnostically relevant populations—particularly goblet cell–containing populations which were confirmed with several downstream analytical techniques.

We identified ECA as an effective lectin for capturing columnar cells, owing to its affinity for β-lactose moieties abundantly expressed in BE and dysplastic tissues. Sugar competition assays confirmed this specificity^40^. While ECA also binds to columnar cells in non-BE contexts such as gastric epithelium, this broader columnar specificity proved advantageous in enriching diagnostically relevant populations, particularly in capsule-sponge samples where BE cells may be sparse. This is further supported by our RT-qPCR data, which showed significantly higher *MUC2* expression in enriched samples, reflecting preferential capture of goblet cell–containing populations characteristic of BE. Future iterations will aim to identify more exclusive markers for distinguishing BE from adjacent columnar epithelium, further enhancing diagnostic precision.

The integration of gold nanoparticles and functionalized PEDOT-pNIPAAM polymer-coated electrodes enhanced cell capture efficiency. The observed improvement in cellular cluster attachment, typically challenging to isolate, underscores the platform’s robustness. The modified polymer surface both captures and relatively quantifies target cells, with successful capture and thermal release demonstrated by EIS data, optical microscopy, and fluorescent staining. Our platform selectively enriches columnar cells even amidst confounding elements like healthy squamous cells or microbial contaminants.

In clinical testing, the platform functions in both static and dynamic configurations. The static setup proved valuable for assessing a single MEA’s sensitivity and specificity and is particularly well-suited for scenarios involving limited cytology samples with lower cell yields, such as fine needle aspirates or brush biopsies. In contrast, the dynamic microfluidic setup enabled efficient enrichment from larger capsule-sponge samples under flow conditions, supporting higher-throughput operation. Both formats allow real-time monitoring and retrieval of viable cells, facilitating downstream genomic and transcriptomic profiling. This adaptability enhances the platform’s clinical utility, offering a cost-effective and portable alternative to conventional EAC profiling workflows^29,30^.

While our results are promising, further validation in larger cohorts is necessary. As an initial proof of concept, we identified limitations for future iterations. Absolute quantification of captured cells on the microfluidic platform proved challenging due to electrode placement differences compared to static measurements. Fabricating planar reference and counter electrodes could address this, though current rapid relative quantification remains beneficial for enrichment applications enabling quick assessment for downstream molecular investigations.

Early detection of pre-malignant lesions is paramount for improving patient outcomes and reducing healthcare costs. In BE progression, precise diagnostic tools are urgently needed. While other approaches typically involve FFPE tissue processing and pathology slides with AI analysis requiring specialized facilities, our bioelectronic platform offers distinct advantages in portability, cost-efficiency, and rapid detection. These features make it particularly suitable for primary care settings or reference laboratories, potentially streamlining diagnosis and improving accessibility to early detection methods. Our study introduces a novel bioelectronic enrichment platform enhancing sensitivity and specificity in detecting BE-associated columnar cells.

While developed for a specific disease context, this technology has potential applications in other malignancies where early detection is critical^41^. The platform’s scalability and adaptability to clinical samples suggest broad application potential including enrichment of circulating tumour cells in liquid biopsies, or even immune or stromal cell populations implicated in early pathogenesis or treatment response^42,43^. By enabling selective enrichment, on-chip relative quantification and downstream analysis of viable cells from minimally invasive samples, our platform sets the stage for a new class of rapid, portable diagnostic tools with broad translational potential across oncology and immunology.

## Methods

### Cell Culture

Human telomerase immortalised fibroblasts (TIF) labelled with red fluorescent protein (RFP–TIF LifeAct) were cultured in DMEM (Thermo Fisher Scientific) supplemented with 20% FBS (Sigma Aldrich), 1% Glutamine (Thermo Fisher Scientific), 2% HEPES (Thermo Fisher Scientific), 1% penicillin–streptomycin (10 000 U mL−1, Thermo Fisher Scientific and 0.1% Gentamicin (Sigma Aldrich). Cells were harvested with 0.25–0.5% trypsin-prior to seeding or passaging. Their viability and counts in all experiments were assessed using the Vi-CELL XR Cell Viability Analyzer (Beckman Coulter).

### Human Tissue

All patients were recruited following informed consent from Addenbrooke’s Hospital (University of Cambridge, UK), with prior peer-review and approval by the ethical committee (Research ethics committee (REC) no. for cohorts 1 and 2 - 07/H0305/52, cohort 3 - 01/149 and cohort 4 - 01/0039) between 2013-2021. Fresh capsule sponge samples were obtained from patients with informed consent at Addenbrooke’s Hospital (University of Cambridge, UK), following ethical approval (REC: 10-H03087-1).

Clinical diagnoses of BE, dysplasia, and cancer were independently conducted by two pathologists based on biopsy samples and BE segment lengths were classified based on endoscopic measurements. Four patient sample cohorts were assembled, each containing formalin fixed paraffin embedded (FFPE) biopsies from individual patients, consisting of BE, GC and NE biopsies. Endoscopic mucosal resection (EMR) samples were included, representing BE, dysplasia, and intramucosal carcinoma (IMC). Furthermore, capsule-sponge slides were used to validate the observations made on biopsy slides in a diverse sampling approach.

### Lectin selection and immunofluorescence determination

Each lectin conjugated with the fluorophore fluorescein isothiocyanate (FITC) was obtained by TCS biosciences, UK at the standard stock solution (1-5mg/mL) which is compatible with IF-based staining. Slides were deparaffinised through xylene and graded ethanol, placed in a humidified incubation chamber, slides blocked for non-specific staining in protein block (Dako Agilent, X0909) for 30min at room temperature, and the FITC-conjugated lectin at its optimised concentration stated above, was applied at 37°C for 15 min on biopsy tissues, and for 30 min on EMR and capsule-sponge tissues. The slides were subsequently washed in lectin buffer and mounted using mounting medium with DAPI (Vectashield plus antifade, 2B Scientific). The lectin-stained slides were imaged the following day, using the ZEISS Axio Scan 7 microscope slide scanner (ZEISS Group, Germany). Two independent pathologists were consulted in evaluating the staining patterns and provide analysis on the tissue specificity and cellular localisation of lectins.

Positive area of staining was quantified to assess the extent of lectin binding pattern. Three fields of view taken at 20x objective per case using the same emission settings, these were then converted into black and white images, and a threshold was set to highlight positive signals in black and background in white, creating a binary image. A common threshold was set for each tissue type based on the tissue sample exhibiting the lowest fluorescent intensity of a positive signal. The positive pixels per area of the field of view were measured and quantified as % total area of positive staining, using the open-source FIJI software^44^. Standard deviation was calculated per sample based on three fields of view, and the mean value was plotted using Python (version 3.10).

### MEA Mask Design and Fabrication

The methodology employed for fabrication of the MEA involves the utilisation of photolithography and the parylene C lift-off method^45^. First, the masks for photolithography were designed which determined all dimensions of the functional features. Two layers of masks were created for each device fabrication using the software Clewin 5.4: a gold-patterning mask and a polymer-patterning mask. The gold patterning mask consisted of the gold features printed in black ink, and the PEDOT:pNIPAAM patterning mask consisted of transparent features for the polymer and contact areas. The final design of the two masks were printed by a micro-photomask printing company, Micro Lithography Services Limited.

The fabrication process followed a previously optimized protocol from our group^46^. It began with cleaning 4-inch glass wafers, followed by photolithography patterning and deposition of a titanium-gold layer using an electron-beam evaporator. After metal lift-off using Technistrip® Ni555, the gold pattern was coated with two layers of parylene using the PDS 2010 Labcoater. Photolithography was then applied to form etching windows for the electrodes, and plasma etching was used to remove the parylene layers in these areas.

### Polymer formulation and spin coating onto MEA

#### PEDOT:PSS synthesis

31 μL of 98% (3-glycidoxypropyl) trimethoxysilane (GOPS) (Merck, UK) was pipetted into 1 mL of a dispersion of PEDOT:PSS (1.3 % wt) (CleviosTM PH1000, Heraeus, Ossila, UK), and then put in an ultrasound bath for 20 mins before use. PEDOT:PSS/pNIPAAm synthesis: 1 mL of a PEDOT:PSS dispersion was mixed with 225 mg pNIPAAm, 15 mg 2,2-dimethoxy-2-phenylacetophenone (DMPA) and 15 mg of N,N’-methylenebis(acrylamide) (mBAAm), all purchased from Merck, UK, with continuous stirring for 30 mins. Then, 31 μL of GOPS was pipetted to the prepolymer mixture and stirred at room temperature for 30mins continuously before use. The molar relationship between PEDOT:PSS and pNIPAAm polymers was calculated to be 1:27.

To create the layers, a similar protocol to the one previously published by our group was followed^47^. Briefly, the desired prepolymer solution was spun for 30 s at 1500 rpm/s, followed by 30s at 3500 rpm/s and baked at 70°C for 2 mins over the electrodes. After that, the layers were photopolymerised using a UV lamp, for 1 min at 365 nm. Then, hardbake was applied to the layers for 2 h at 120°C. Once the polymerisation was complete, the layers were rinsed with ethanol and DI water to eliminate the non-polymerised monomers, resulting in a thin film of the polymeric layer on the substrate.

### Lectin Functionalisation on polymer-coated MEAs

Lectin functionalisation on the bespoke MEA platform was achieved via covalent coupling onto AuNPs. The process began with plasma treating polymer-coated MEAs (100 W, 2 min, Air) to introduce hydrophilicity. Then, 50μL of 0.1M Poly-L-Lysine (PLL) (Merck, USA) was applied and incubated for 30 minutes to induce a net positive charge on the surface. Next, 50nm lipoic-acid coated AuNPs (50μg/mL, Nanocomposix, UK) were loaded onto the PLL-coated MEAs for 4 hours to allow electrostatic binding. EDC and sulfo-NHS (Sigma Aldrich) were mixed at a 1:2 ratio in PBS (pH 7.4) and applied to the AuNP-coated surface for 30 minutes at room temperature. Immediately after, the surface was washed with PBS and loaded with 50μg/mL FITC-Lectin ECA for overnight incubation. Functionalization was confirmed via fluorescence microscopy and EIS measurements.

### AFM/SEM characterisation

The polymer PEDOT:pNIPAAM was spin-coated onto a glass cover slip based on the protocol stated above and used for AFM-based characterisation. The experiments were carried out in Scanasyst tapping mode using ScanasystFluid+ probes (Bruker, USA) with a nominal spring constant of 0.7 N m–1 and a resonant frequency of 150 kHz. Images were recorded at scan speeds of 1.5 Hz and tip–sample interaction forces between 200 and 300pN. To resolve the morphology of the polymer layers, 5 × 5 μm scans were generated. Scans were taken of the polymer at room temperature first, which were then heated to 37°C and scanned again, to evaluate the modified surface morphology. Measurements of the peak height of the polymers were performed by taking cross-sections across different areas of interest using the Nanoscope analysis software (Bruker, USA).

### Microscopy-based optical characterisation

Brightfield and fluorescence microscopy was carried out as part of the optical characterisation of cell capture and release on different lectin-functionalised polymer substrates. All experiments were carried out using the Zeiss Axio Observer with the 20×/0.8, (Plan-Apochromat, Zeiss) objective. The brightfield, EGFP (488nm), DAPI (359nm) and RFP (588nm) channels were used in most experiments together with fixed laser intensities.

### Microfluidic-based dynamic cell enrichment

After lectin functionalisation and parylene lift-off, a 60μm-height microfluidic channel with an adhesive bottom (μ-Slide I Luer, IBIDI, Martinsried, Germany) was manually aligned and placed onto a rectangular chip containing 4 MEAs, ensuring that the 5mm width of the channel covered all 6 electrodes of each MEA.

Cellular sample processing was performed using a syringe pump (Harvard Apparatus Pump 33 DDS, USA) with reverse flow. The sample inlet on the right of the channel was connected to diluted cellular samples via the Luer inlet, while the outlet on the left was attached to a 20mL syringe (BD Plastipak™ with BD Luer-Lok™, Switzerland). Flow rates were optimized between 20-50μL/min, with lower rates used in the first hour to enhance cell capture. After initial processing and EIS measurements, flow rates were increased for efficient sample processing.

### Electrochemical measurements

All electrochemical measurements were performed using a PalmSens4, equipped with PSTrace 5.6 software (PalmSens BV, Houten, The Netherlands). A three-electrode system was employed during all EIS measurements. The platinum served as an auxiliary electrode and Ag/AgCl served as a reference electrode in 1X PBS electrolyte containing 5 mM [Fe (CN)6]^3−/4−^ and 0.1 M potassium chloride (KCl) at a 0.01 V potential over the frequency range from 100 kHz to 1 Hz.

EIS measurements were carried out on the polymer-coated MEA platform using a fixed plastic well covering the 6 electrode areas. Here, the contact pads designed with a specific spacing was used to align with that of the pogo pins to make contact across the electrodes in the MEA at once. However, each electrode was queried separately to obtain independent electrode measurements. For EIS measurements carried out on MEAs within the fluidic channel, the PBS electrolyte containing 5 mM [Fe(CN)6]^3−/4−^ was passed through the channel to completely cover all the MEAs along with the inlet/outlet. The two MEAs placed in the middle of the channel were queried by EIS. The RE and the CE were dipped on either the inlet or the outlet, depending on its proximity to the MEA being measured. The pogo pins were used similarly as stated above to make contact with the exposed contact pads to act as the WE and complete the circuit for EIS measurements. Impedance spectra were fitted to a simplified Randles equivalent circuit consisting of solution resistance (Rs), double-layer capacitance (Cdl), and charge-transfer resistance (Rct). Curve fitting was performed using PSTrace 5.6 and Rct values were extracted for quantitative comparison

### Clinical Validation Experiments

#### Cytospin™ of Released Cells

Cell aliquots from the clinical validation experiment were collected, kept on ice, centrifuged at 400 x g (4°C) for 5 mins, and washed with 1mL ice-cold PBS. After centrifugation, cells were fixed with 500μL of 4% paraformaldehyde (PFA) for 20 mins on ice, then centrifuged again, washed, and resuspended in PBS. Fixed cells were transferred onto SuperFrost Ultra Plus™ adhesion slides (Thermo Fisher Scientific, USA) using a Cytospin 3 centrifuge (Shandon Southern Instruments, USA) at 500rpm for 5 mins. Slides were dried overnight at room temperature (22°C) in the dark.

#### H&E Staining of Cytospin Slides

Slides was stained in a Bond autostainer (company, country) with haematoxylin (Gill’s II, VWR, USA, code: 1.05175.0500) for 2 mins, followed by eosin staining (1% eosin, VWR code: 34197) with 1% calcium carbonate (Sigma-Aldrich, Merck, USA, code: C4830). After washing, the slides were dehydrated with ethanol series (70%, 95%, 100%) and cleared in Histoclear before being mounted with coverslips.

#### RT-qPCR Evaluation of RNA from Clinical Samples

RNA extraction was performed using the Qiagen AllPrep DNA/RNA Micro kit (Qiagen UK, Cat. No. 80284), and cDNA was synthesized using the Qiagen QuantiTect® Reverse Transcription kit (Qiagen UK, Cat. No. 205311). RT-qPCR was carried out with the Qiagen QuantiNova Probe PCR kit (Qiagen UK, Cat. No. 208252). Technical replicates were conducted for genes *MUC5AC, MUC2* with *GAPDH* and *β2M* as housekeeping genes. Relative gene expression was calculated using the ΔΔCT method. Sequences for the primer sets used are stated in ESI (ESI, Table 3).

## Author contributions

S.P. and R.C.F. conceptualised the whole study. S.P. led the platform integration, electrochemical analysis, data interpretation and manuscript preparation. N.M.-S. conducted initial lectin staining and characterisation experiments and helped conceptualise lectin biomarker validation experiments. Z.L. microfabricated the electrode arrays. Y.F. co-developed the hybrid polymer and supported characterisation.

J.S. optimised the polymer formulation and supported imaging. A.U. conducted patient sample staining and validation. A.B.P. contributed to SEM analysis and nanoparticle-polymer integration. J.L.Z. assisted with qPCR experiments. I.M. helped to acquire and analyse AFM data. L.F. and R.M.O. provided project guidance and technical input. The project was supervised by R.M.O, L.F and R.C.F.

## Acknowledgements

This work was supported by the Medical Research Council [MR/W014122/1] and Rosetrees CIO/Stoneygate Trust [CF-2021-2\136]. The Human Research Tissue Bank is supported by the NIHR Cambridge Biomedical Research Centre (NIHR203312). L.F. acknowledges funding from the EPSRC Interdisciplinary Research Centre grant (EPSRC IRC EP/S009000/1). S.P. acknowledges support from the Cambridge Commonwealth, European & International Trust, Trinity College at the University of Cambridge, and the EPSRC Centre for Doctoral Training in Sensor Technologies and Applications (EP/L015889/1). Graphics shown in Figures 1–4 were created with BioRender.com.

## Data availability

All data supporting the findings of this study are available within the article and its supplementary information files. Additional data are available from the corresponding author upon reasonable request.

