## Supplementary Information for "Bioelectronic platform enables lectin-based enrichment of columnar cell clusters for Barrett’s oesophagus detection"

### Electronic Supplementary Information:

**Table ESI-1. Categories and concentrations of lectins tested in the discovery study.** Four classes of lectins with different glycan-binding domains were tested to cover the landscape of altered landscape of cell-surface glycans in gastro-oesophageal tissues in normal and disease states. The concentrations tested were within the range stated by the manufacturer. Concentrations stated in bold indicate the final optimised concentration used, where contextual tissue specificity was observed within the tissue types being tested (i.e NE, GC or BE).

| Glycan Binding Specificity | Lectin | Concentrations Tested (μg/ml) |
|---|---|---|
| O-Glycan Binding (LacNac, GalNAc binding) | *Erythrina cristagalli lectin* (ECA) | 1, 3, 5, 10, **20**, 40 |
| | *Griffonia simplicifolia lectin I* (GS-I) | 1, 5, **20** |
| | *Maackia amurensis lectin I* (MAL-I) | 1, 5, **20** |
| | *Vicia villosa agglutinin* (VVA) | 1, 5, **20** |
| | *Peanut agglutinin* (PNA) | 1, 3, 5, 10, **20** |
| | *Ricinus communis agglutinin I* (RCA-I) | 1, 5, **20** |
| | *Glycine max soybean agglutinin* (SBA) | 1, 5, **20** |
| | *Dolichos biflorus agglutinin* (DBA) | 1, 5, **20** |
| | *Cytisus scoparius* (CSA) | 1, 3, 5, 10, **20** |
| | *Helix pomatia agglutinin* (HPA) | 1, 5, **20** |
| | *Bauhinia purpurea agglutinin A* (BPA) | 1, 5, **20** |
| Sialic Acid Binding | *Triticum vulgaris wheat germ agglutinin* (WGA) | 1, 3, 5, **20** |
| | *Sambucus nigra agglutinin I* (SNA-1) | 1, 5, **20** |
| | *Artocarpus integrifolia lectin* (Jacalin) | 1, 5, **20** |
| | *Amoora cucullata agglutinin* (ACA) | 1, 5, **20** |
| Fucose Binding | *Ulex europaeus agglutinin I* (UEA-1) | 1, 5, **20** |
| | *Aleuria aurantia lectin* (AAL) | 1, 5, 10, **20** |
| | *Salvia japonica agglutinin* (SJA) | 1, 5, 10 |
| Complex N Glycan | *Phaseolus vulgaris erythroagglutinin* (PHA-E) | 1, 3, 5, **20** |
| | *Concanavalin A* (Con A) | 1, 5, **20** |

**Table ESI-2. Summary of lectin staining pattern on patient biopsy tissue.** Red indicates a complete lack of staining, while green indicates an observed positive staining pattern. Specific cellular localisation in each positively stained tissues are described in their respective boxes.

| Lectin | Sugar Specificity | Sugar Structure | Tissue Specificity | | |
|---|---|---|---|---|---|
| | | | NE | GC | BE |
| WGA | N-glycan (GlcNac) with sialic acid | 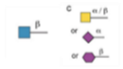 | Membrane | | Goblet +ve |
| Jacalin | T-antigen (mono or di sialated) (Gal (β-1,3)GlcNac) | 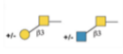 | Membrane | | Goblet +ve |
| RCA-1 | Galactose, Lactose | 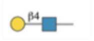 | Membrane | | Epithelial |
| UEA-1 | L-fucose | 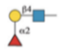 | Membrane | | Epithelial |
| AAL | L-fucose | 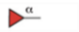 | Membrane | | Goblet +ve |
| PHA-E | N-glycan with Gal-(GlcNAc) | 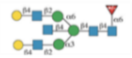 | Membrane | | |
| SJA | Gal-GalNAc<br>Mannose, Glucose | 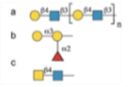 | | | |
| BPA | Galβ3GalNAc | 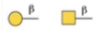 | | | |
| HPA | α-GalNac | 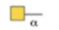 | | | |
| SNA-1 | Sialic Acid- Gal/ GalNac | 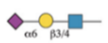 | | | |
| Con A | Mannose, Glucose | 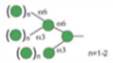 | | | |

| Lectin | Sugar Specificity | Sugar Structure | Tissue Specificity | | |
|---|---|---|---|---|---|
| | | | NE | GC | BE |
| ECA | Lactose | 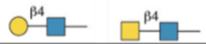 | | Foveolar +Mucin | Goblet +ve |
| GS-1 | Galactose, GalNac | 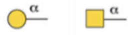 | | Foveolar +Mucin | Deep glands |
| MAL-1 | Lactose | 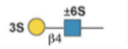 | | Foveolar +Mucin | Deep glands |
| SBA | Terminal α- or β-linked GalNac | 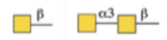 | | Foveolar +Mucin | Mucin cap |
| DBA | α-linked GalNac | 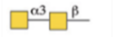 | | Mucin | Mucin cap |
| PNA | T-antigen (un-sialated only) (Gal (β-1,3)GlcNac) | 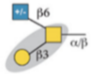 | | Foveolar +Mucin | |
| CSA | GalNac | 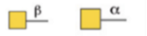 | | Foveolar +Mucin | |
| ACA | Galβ3GalNAc Sialic acid | 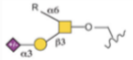 | | Foveolar +Mucin | |
| VVA | α- or β-linked terminal GalNac | 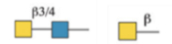 | | Foveolar +Mucin | |

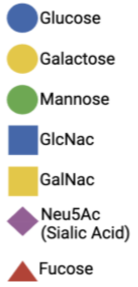

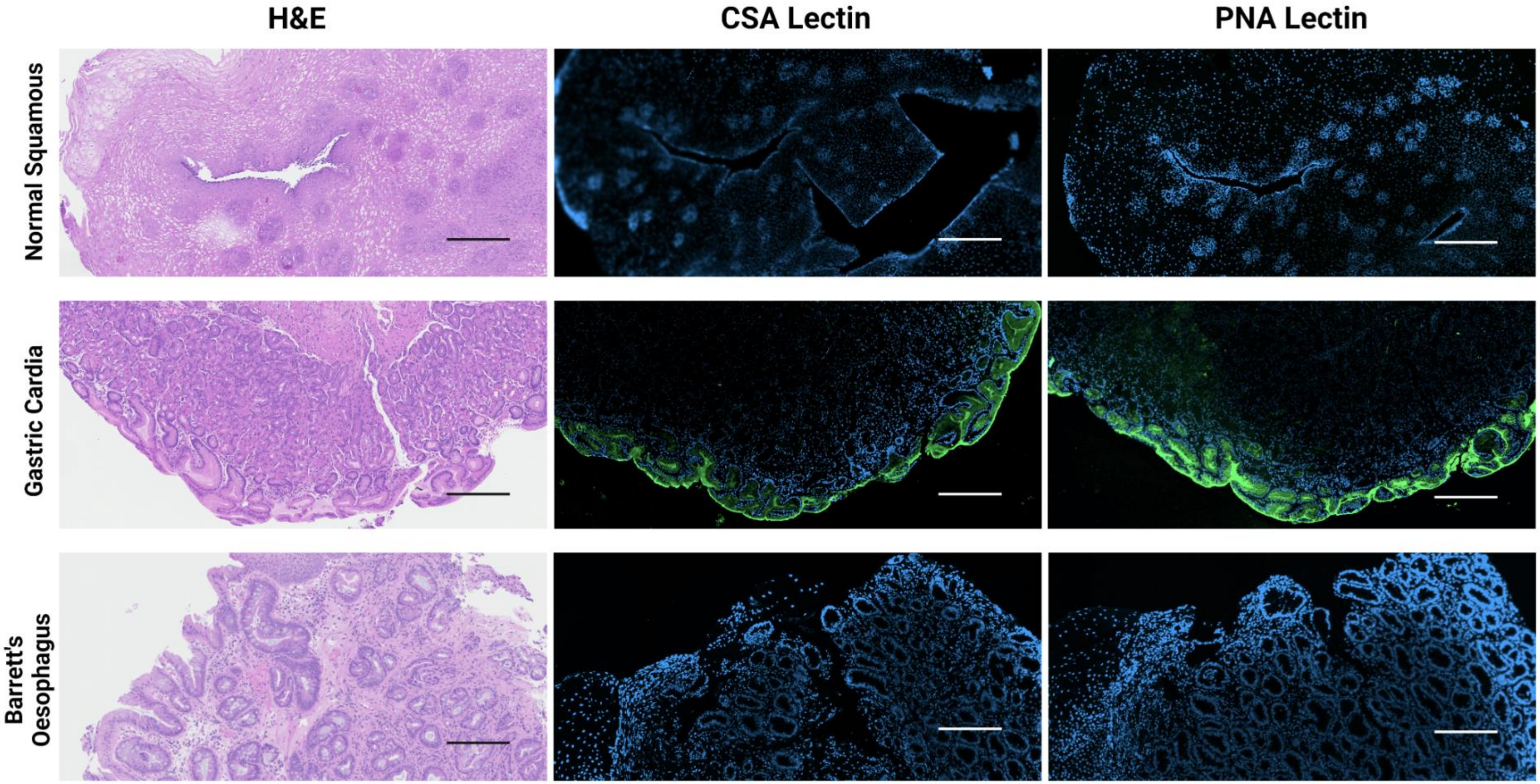


**Figure ESI-1. Representative images of adjacent H&E, and lectins CSA and PNA staining of biopsy trios.** Specific staining of GC tissue was observed for both lectins CSA and PNA. Scale bars for all tissues indicate 200μm.

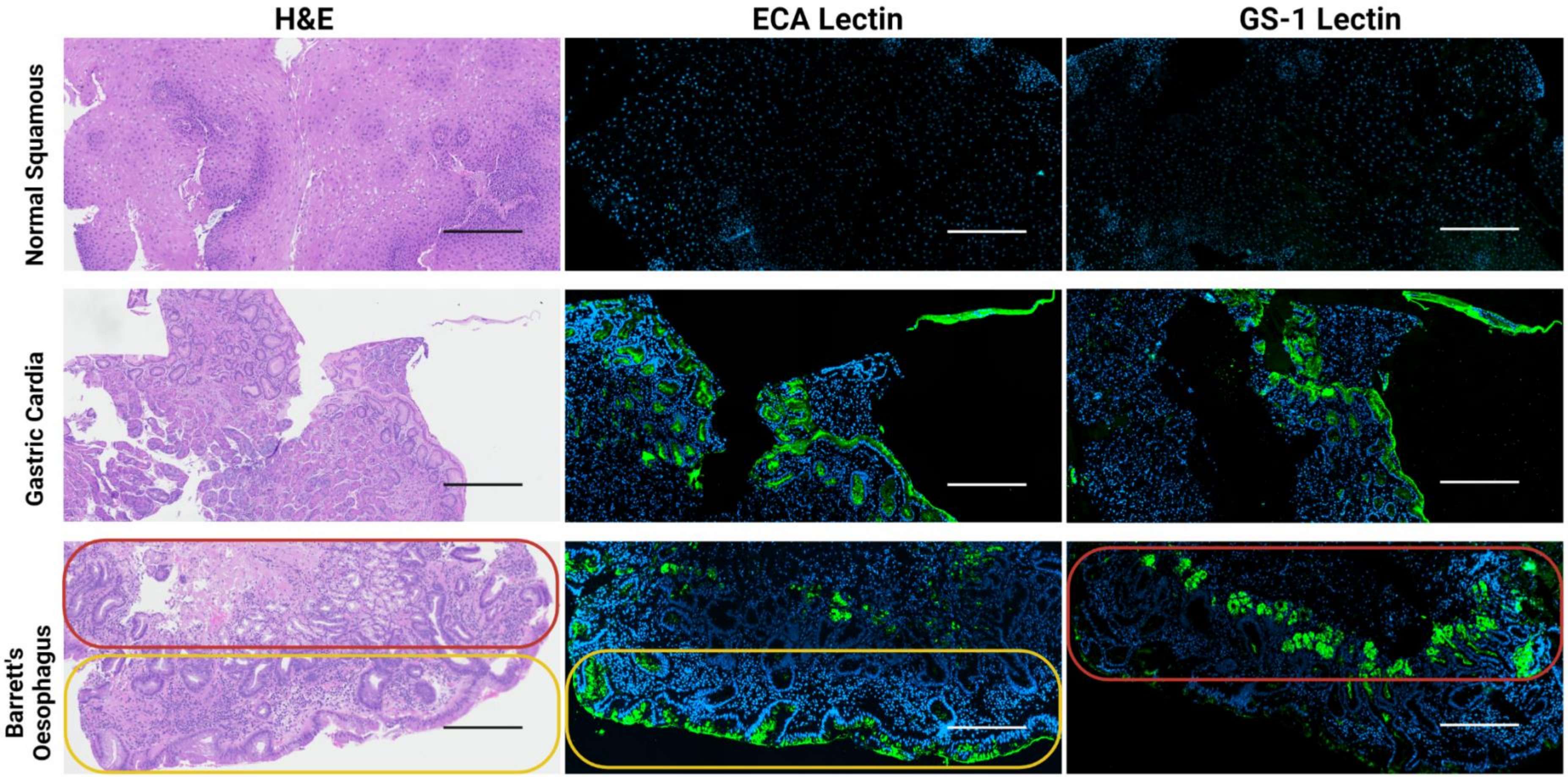


**Figure ESI-2. Representative images of adjacent H&E, and lectins ECA and GS-1 staining of biopsy trios.** Specific staining of GC and BE tissue were observed for lectins ECA, GS-1. Scale bars for all tissues indicate 200μm. Superficial glands present at the epithelial surface (yellow region), along with glands located deeper in the tissue (red region), are highlighted

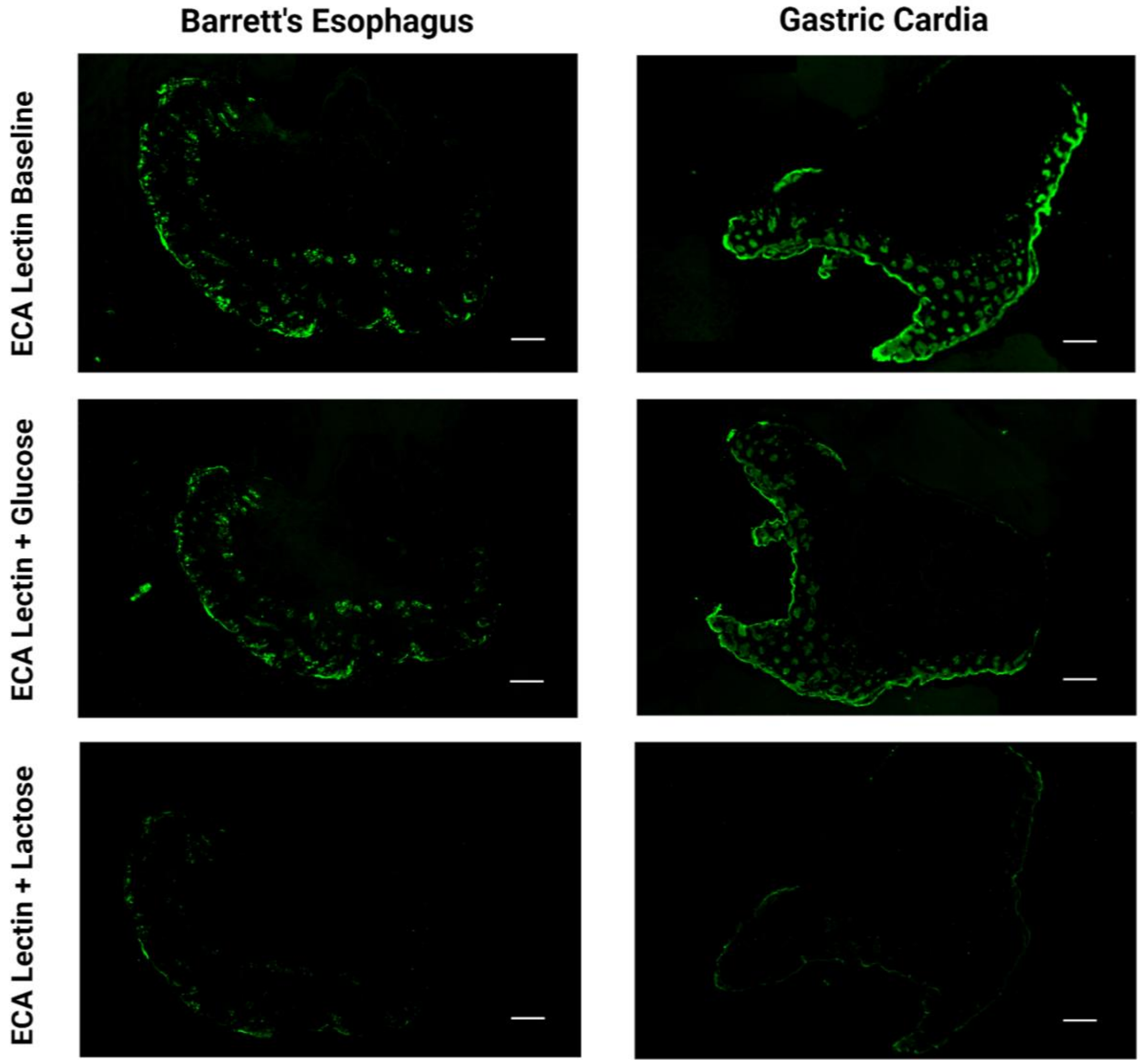


**Figure ESI-3. Representative images from the ECA lectin - sugar competition assay in biopsy specimen (AHM1394).** Depicts the loss in positive staining for D-lactose mixed in sample. Scale bars are 200µm (n=3 for each condition).

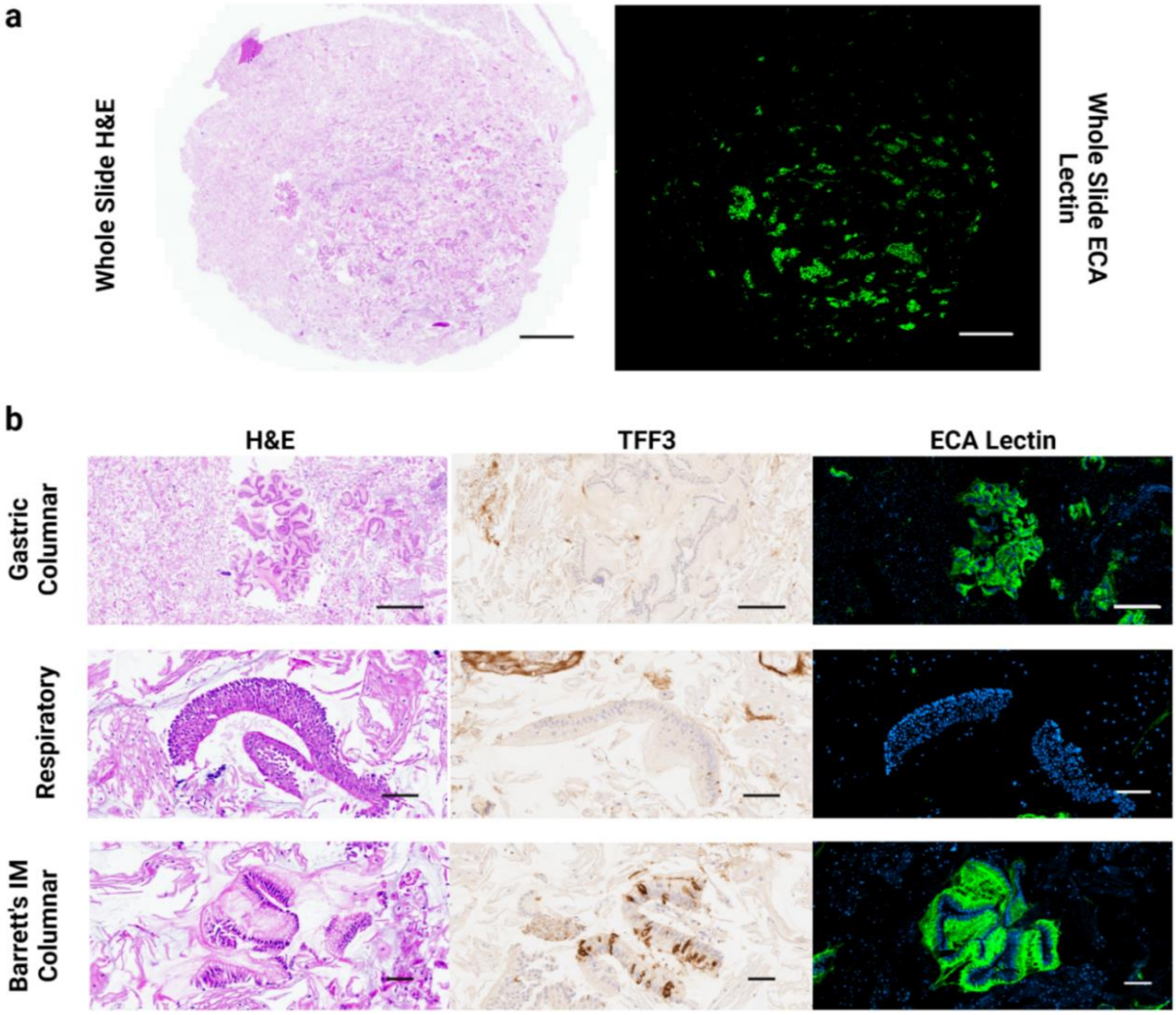


**Figure ESI-4. Candidate lectin ECA validation study in Cytosponge sampling.** a) Adjacent whole slide images of H&E and ECA lectin IF stained Cytosponge cellular sample (BEST2/UCL/0228) with scale bars – 2mm. b) Magnified regions from the whole slide depicting different cell types in the sampling (gastric columnar, respiratory, and Barrett's IM columnar). Adjacent images of H&E, TFF3 and ECA lectin staining in each cell type. Scale bars – 100μm.

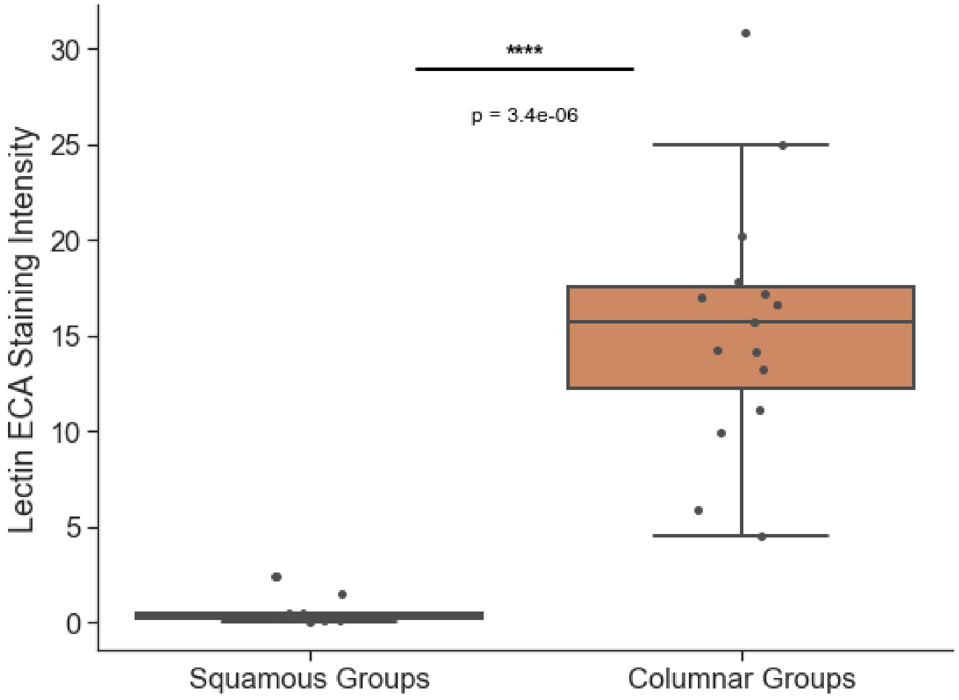


**Figure ESI-5. Quantified % area of positive ECA staining in Cytosponge samples across different fields of view for each cell group in cohort 4.** Three fields of view were used for each group within a sample (n=5). Independent t-test, **** p ≤0.0001. Specific staining for columnar groups was observed consistently across all Cytosponge samples.

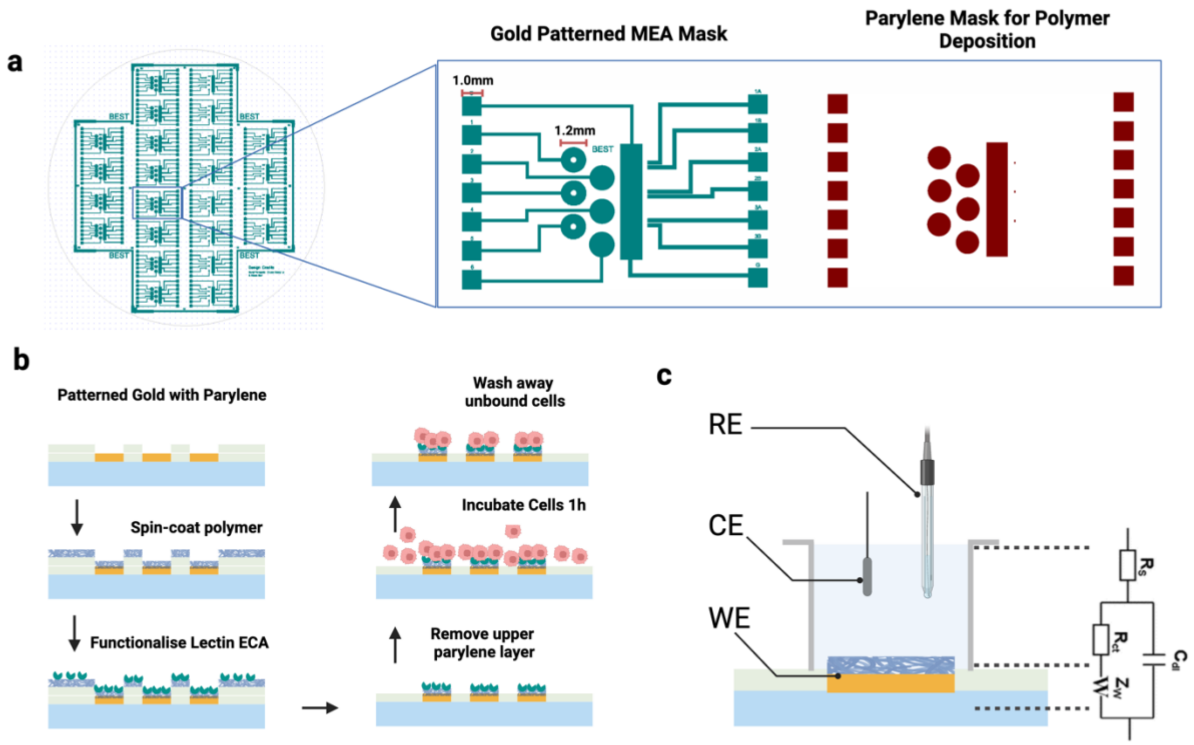


**Figure ESI-6. Bespoke MEA design for bioelectronic enrichment.** a) The 4-inch wafer design consisting of multiple MEAs. Inset represents the fabrication masks for gold (left) and parylene (right) patterning. b) Schematic depicting the strategy employed for localised spin-coating of the hybrid polymer and localised functionalisation of lectins. c) The electrochemical configuration used for EIS measurements. RE - Reference Electrode (Ag-AgCl), CE – Counter Electrode (Pt), WE – Working Electrode (PEDOT-pNIPAAM). Randle's circuit was used to fit and quantify the EIS measurements.

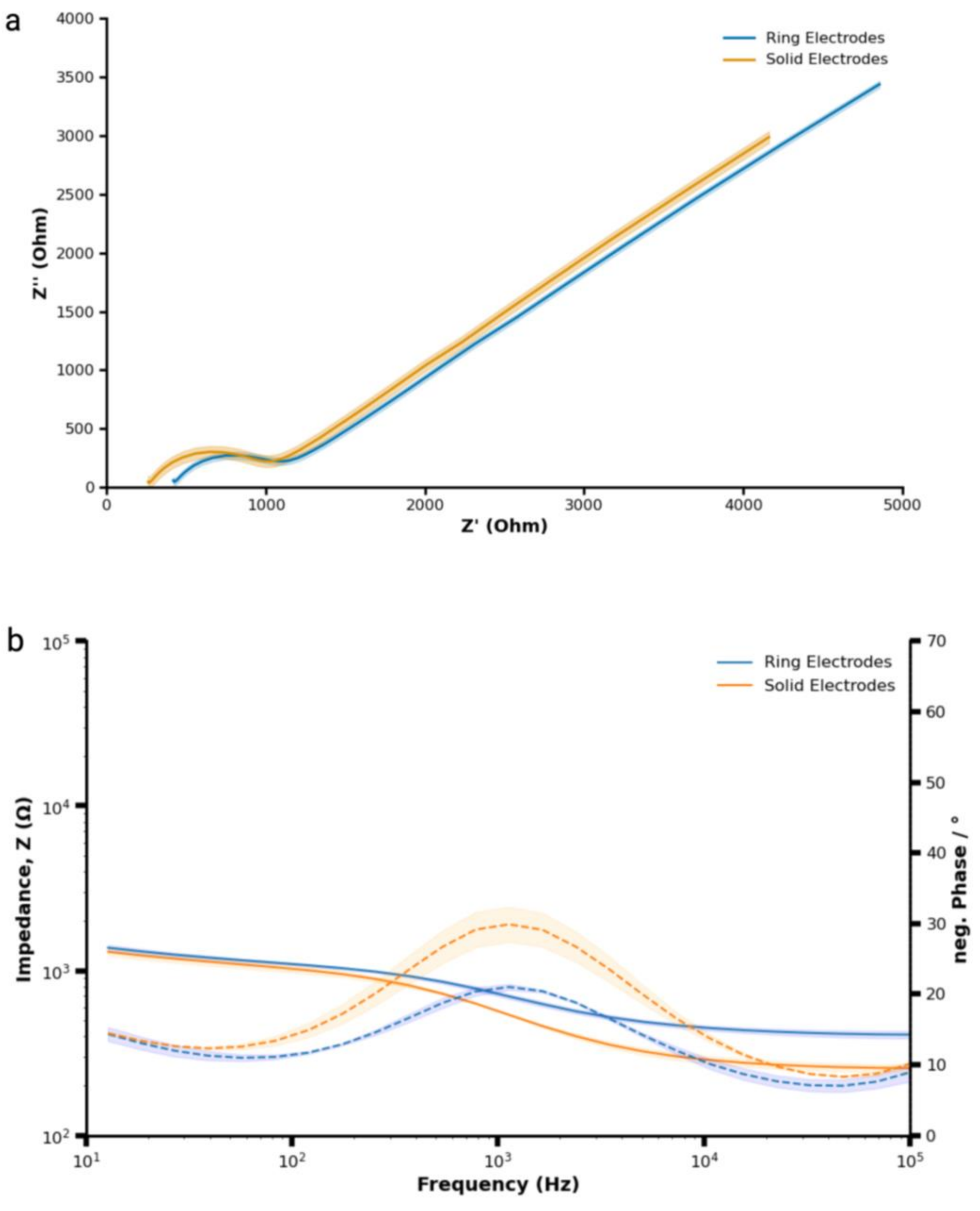


**Figure ESI-7. Baseline EIS measurements of fabricated bare gold electrodes comparing the two types used in the MEA.** a) Nyquist and b) Bode plots demonstrating the baseline consistency between ring electrodes (n=3) and the standard circular electrodes (n=3). The shading represents the standard deviation of the mean for 3 electrodes. In the Bode plot, the left axis represents magnitude (solid lines), and the right axis represents phase (dashed lines).

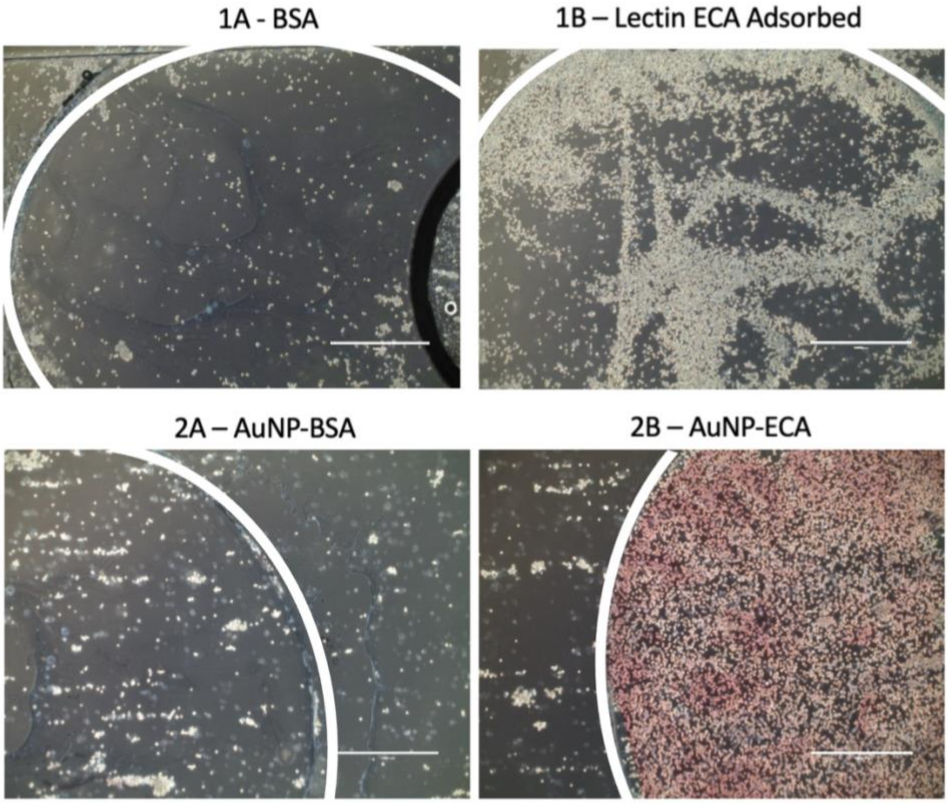


**Figure ESI-8. Comparison of dynamic microfluidic cell capture based on direct lectin adsorption vs AuNP-based functionalisation.** All figures represent a glass slide coated with the polymer PEDOT-pNIPAAM using a 1.2mm diameter mask consistent with the electrode area in the MEA. The white circular arcs demarcate the functionalised area, to visualise the cell capture (TIF-LifeAct RFP) region across all conditions. 1A, 1B depicts dynamic cell capture in a microfluidic channel, using a direct adsorption method where either BSA (1A) or lectin ECA (1B) was adsorbed onto the polymer coated area on the slide. 2A, 2B represents the cell capture using the AuNP functionalisation approach presented in Fig. 3. A homogenous capture of cells was observed for the AuNP-functionalised region, compared to sparse/ heterogenous capture of cells seen for areas directly adsorbed with lectin ECA. Scale bars are 500µm. The characteristic reddish-pink appearance in panel 2B could result from the collective optical properties of surface-bound AuNPs stabilized through specific cell-lectin interactions. In contrast, the absence of color in the AuNP-BSA control (2A) suggests that non-specific interactions lead to AuNP dispersion and removal during cell addition and washing steps.

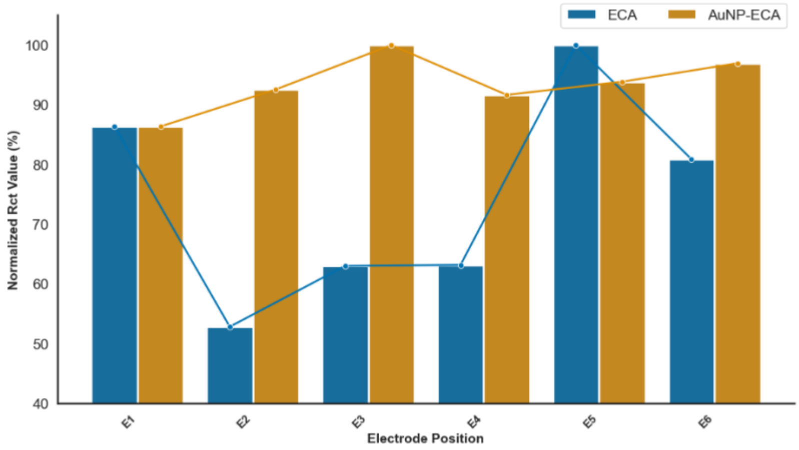


**Figure ESI-9. Lectin functionalised baselines for direct lectin adsorption vs. AuNP-based functionalisation**. Comparison of normalised EIS quantification data for lectin functionalised baselines for direct adsorption versus AuNP-based attachment. A relatively large variation was observed for directly adsorbed lectins on the MEA, indicating a sparse or patchy coating. In comparison, the AuNP-functionalised ECA baselines demonstrated a more consisted Rct distribution across the 6 electrodes.

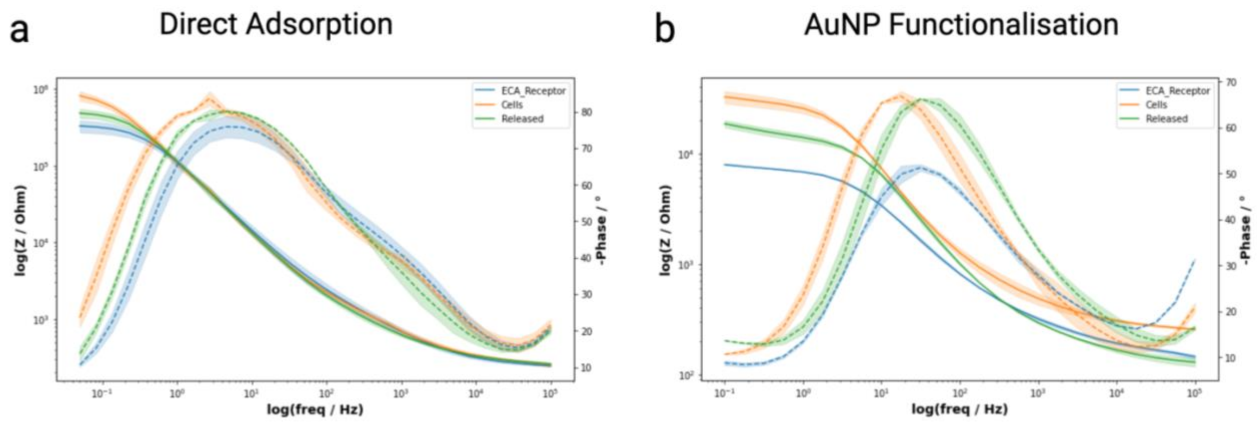


**Figure ESI-10. EIS measurement sensitivity for direct lectin adsorption vs. AuNP-based functionalisation**. Comparative Bode plots of cell capture and release data for direct adsorption of lectins (a) versus AuNP-based functionalisation. The plots present depict an enhanced sensitivity in detecting surface binding/release events with AuNP-functionalised electrodes.

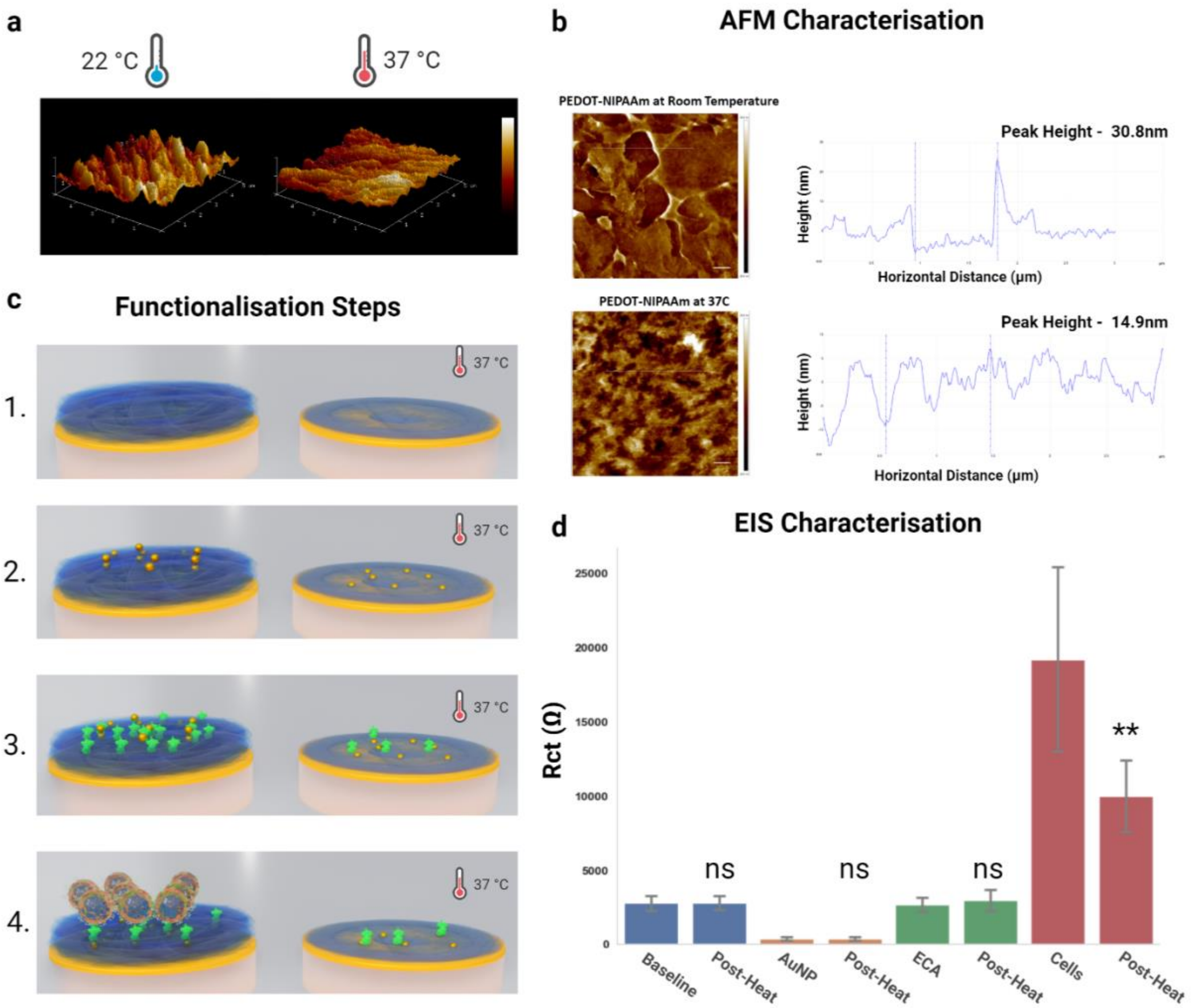


**Figure ESI-11. Mechanistic studies to evaluate the mechanism of cell release from the platform.** a) 3D rendering of AFM data depicting the surface morphology of the polymer PEDOT:pNIPAAM at room temperature (RT) and post heating to 37°C. b) Representative 2D AFM with scale bar - 1µm and line plots of the polymer indicating the vertical heights of the polymeric chains at RT and at 37°C. c) Graphical depiction of the mechanistic characterisation experiment. d) Circuit-based quantification of Rct from the EIS measurements taken at each step during the experiment, starting with the polymer baseline. Error bars correspond to the standard deviation from the mean for n=18. Pairwise t-test with Bonferroni correction between the RT and post-heat condition for each step was carried out. Only the post-heat condition after cell capture indicated a significant difference *** $p \leq 0.001$, indicating that previous functionalisation layers of AuNPs and lectins remained mostly the same post heat treatment.

a

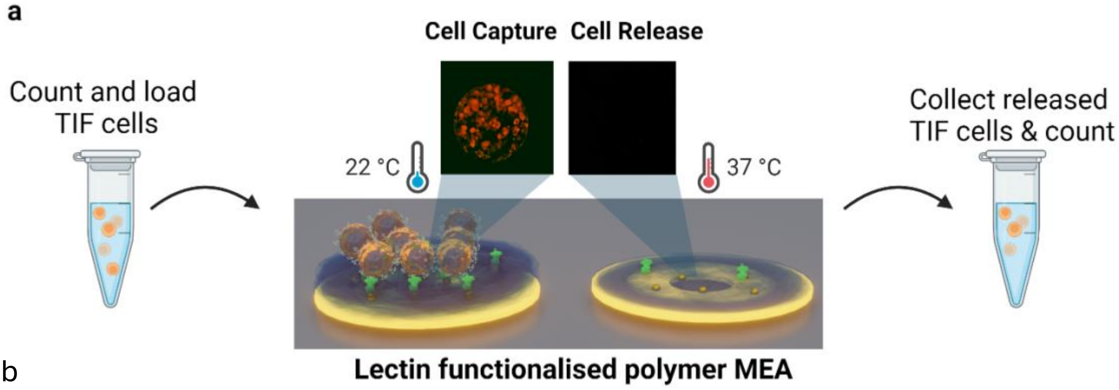


b

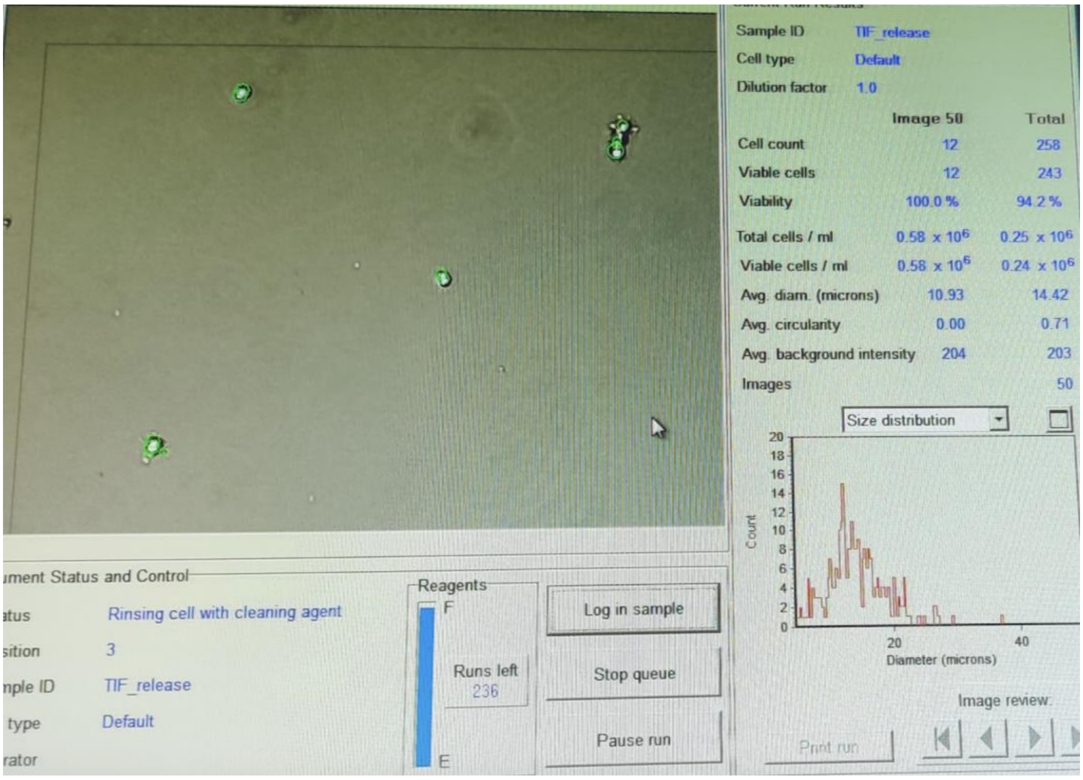


**Figure ESI-12. Thermal Detachment Efficiency.** a) Schematic of experiment for assessing numbers and viability of cells released from the platform. b) ViCell cell counter data on cell retrieval efficiency and viability post-thermal detachment, confirming a retrieval rate of 78% efficiency and 95% viability(n=3). The density of retrieved cells was small (hence the sparse field of view, FOV).

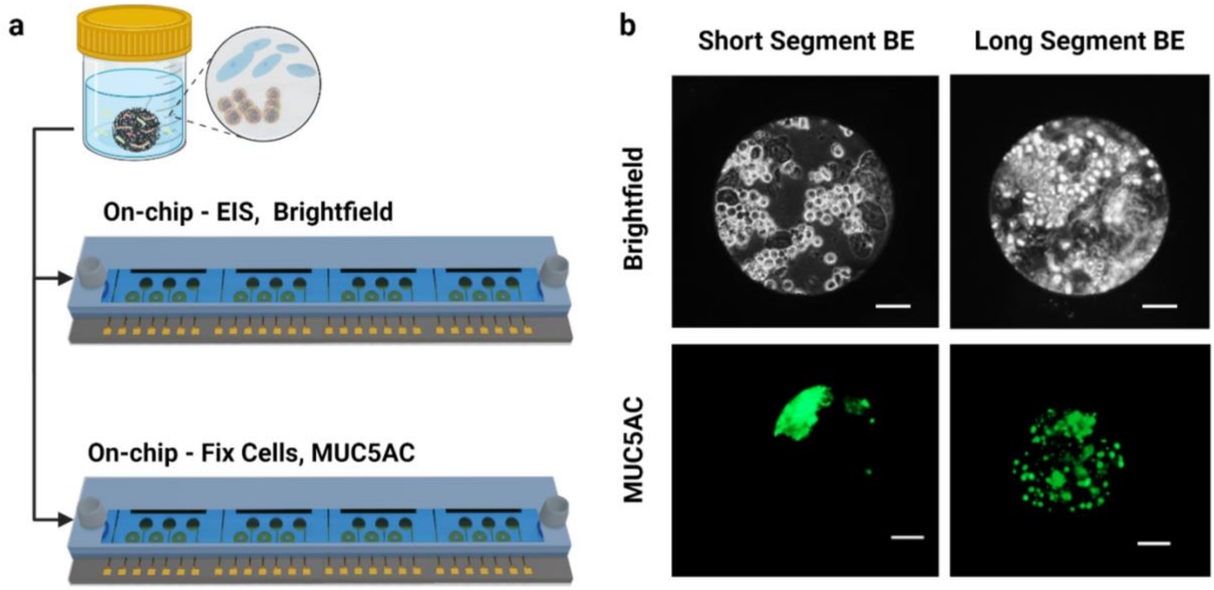


**Figure ESI-13. Optical characterisation of cells captured within the electrode's optical window during Cytosponge sample processing of BE cases.** a) Graphical representation of two lectin-functionalised fluidic MEA devices, wherein the primary device (top) was used for brightfield evaluation of cell capture, and secondary device (bottom) used for on-chip immunofluorescence evaluation. b) MUC5AC IF staining carried out on cells captured in a parallel microfluidic MEA device. Scale bars are 100µm. These results help verify the identity of the captured cellular clusters on the electrodes to be columnar groups.

**Table ESI-**Error! No text of specified style in document.. **Primer sets for RT-qPCR studies.**

| | |
|---|---|
| MUC2_forward | GCCGGCTATTACCACACAGA |
| MUC2_reverse | TGTTATGGACGCAAGGGCAT |
| MUC5AC_forward | TATGTGCTGACCAAGCCCTG |
| MUC5AC_reverse | TTGATCACCACCACCGTCTG |
